# Social and Psychiatric Effects of COVID-19 Pandemic and Distance Learning On High School Students: A Cross-Sectional Web-Based Survey Comparing Turkey and Denmark

**DOI:** 10.1101/2020.10.21.20217406

**Authors:** LS Seyahi, SG Ozcan, N Sut, A Mayer, BC Poyraz

**Affiliations:** Senior High School Student, American Robert College, Istanbul, Turkey; Cerrahpasa Medical Faculty, Department of Internal Medicine, Cerrahpasa Medical School, Istanbul University-Cerrahpasa, Istanbul, Turkey; Department of Biostatistics and Medical Informatics, Trakya University Medical Faculty, Edirne, Turkey; Senior High School Student, Aurehøj High School, Copenhagen, Denmark; Department of Psychiatry, Division of Geriatric Psychiatry, Cerrahpasa Medical Faculty, Istanbul University-Cerrahpasa, Istanbul, Turkey

**Keywords:** COVID-19, education, adolescents, distance learning, psychology, sleep problems, eating problems, domestic abuse, positive and negative affect, PANAS, web based survey, Turkey, Denmark

## Abstract

**Objective:** In this study we investigated the socio-psychological effects of both the pandemic and distance learning on high school students in Turkey and Denmark. We aimed to assess whether there were any differences a) between students attending public or private schools in Turkey and b) between two countries having different approaches to pandemic and considerable socio-cultural and economic differences.

**Methods:** We conducted a web-based questionnaire study in a cross-sectional design using Survey Monkey platform and sent out via social media to high school students from Turkey and Denmark. The survey collected socio-demographic data, several variables associated with pandemic and distance education and their effects on social life and psychological status. Additionally, emotional status was assessed using positive (PA) and negative affects (NA) schedule (PANAS). The survey ran from July 3 and August 31 2020.

**Results:** We studied 565 (mean age: 16.5 ± 1.0) Turkish and 92 (mean age:17.7 ± 1.0) Danish students, of whom the majority were female adolescents (63% vs 76%). Students educated in public (47.6%) and private high schools (52.4%) were nearly similar in number in Turkish group, whereas in the Danish sample almost all students were from public school (98.9%). Turkish students were significantly more likely to be compliant with the pandemic related restrictions. Besides that, there were significant socio-economic disparities between Turkish and Danish students and also within Turkey between public and private school students. Turkish online education system was significantly less adequate and satisfactory compared to the Danish system. These were even worse for those who were attending public schools in Turkey. Regardless of the socio-economic differences, the majority of the students in both countries has been negatively affected by the pandemic and related restrictions and had a negative opinion about distance education. This was also true for the PANAS scores. The total scores of PANAS were similar between Turkish and Danish students (PA: 27.0 ± 7.6 versus 25.8 ± 5.6; NA: 24.8± 7.5 versus 24.5± 7.3) and also within Turkey between public and private school students (PA: 26.8 ± 7.5 versus 27.1 ± 7.6; NA: 24.7± 7.2 versus 25.0± 7.8). While female students were significantly more severely affected in the Turkish group, no such gender differences were observed in the Danish group. Additionally, considerable portion of the students in Turkey and Denmark expressed loneliness (55.2% vs 59.8%, p<0.706), boredom (71.2% vs 58.7%, p=0.019) and anxiety towards the future (61.4% vs 22.8%, p<0.001). Decreased physical activity, sleep problems, eating disorders and domestic abuse were other complaints.

**Conclusions:** Adolescents from both countries have been severely affected by the pandemic and its related restrictions and expressed negative views about distance education. Turkish online education system seemed to be less satisfactory when compared to Danish system and within Turkey, public school students had significantly more disadvantages compared to those attending private schools. Despite the fact that there were several socio-economic inequalities among students, in general, there were no robust significant differences regarding psychological status and opinion about distance learning, indicating a global worsening of emotional status during pandemic.

## INTRODUCTION

On March 11 2020, The World Health Organization declared the pandemic status of a new type of coronavirus (severe acute respiratory syndrome coronavirus 2 [SARS-CoV-2]) infection (COVID-19) (1). Many countries around the globe responded with unprecedented public health measures to control the spread of the infection. The closure of the schools and switching to online education had a considerable impact on the daily life of the adolescents. We hypothesized that the impact would be greater among economically disadvantaged adolescents. While current data indicate that adolescents’ psychological status and motivation are negatively affected by the pandemic and online education (2-10), to the best of our knowledge, data on cross-cultural differences and whether there are any differences among public versus private school students are largely missing. Therefore, in this study we aimed to analyze the effects of both the pandemic and distance learning on social and psychological status of high school students in Turkey, with special emphasis to see whether attending public or private school systems had an effect. Also as a control group we studied high school students from Denmark, an European Union country with significant socio-cultural and economical differences and a different approach to pandemic compared to Turkey.

## METHODS

### Study Design

#### a) Identification of the participants

The survey was sent out to high school students (Grades 9, 10, 11 and 12 for Turkey, Grades 9, 10, and 11 for Denmark) via WhatsApp and Instagram. Snowball sampling was used by asking students to send the survey link to their peers via their social media. We did not make a sample size analysis. However, as one of the study objectives was to compare public and private schools, in Turkey, we intended to reach at least 100 students each attending public and private schools.

#### b) Country-specific pandemic control measures and distance education systems

##### 1) Turkey

The first case of COVID-19 in Turkey was identified on March 11, 2020 (11) Turkish Government issued several regulations to control the spread of the virus. All schools were closed as of March 16 (12). The closure was eventually extended to continue until the school year ended. In April, 2020, a total curfew was declared for people younger than 20 years of age until early June. As of September 8, mask wearing became mandatory in all public areas (13).

Turkey shifted into distance learning in mid-March. During this time, the Ministry of Education led distance learning through the Education Informatics Network (known as EBA in Turkey) using television broadcasts and online lessons on their websites of which were no more than 2 hours/day (14). The education in public schools became heavily dependent on the EBA, compared to this; private schools (and certain more resourceful public schools) continued their curriculum through their own systems with more frequent classes and more technological equipment.

##### 2) Denmark

The first case of COVID-19 was identified in Denmark on February 27, 2020 (15). Denmark was one of the European countries that introduced lockdown measures at the earliest. In mid-March, primary schools, universities, libraries, indoor cultural institutions and similar places were closed. A month later, the government embarked on its gradual reopening by letting the youngest children goes back to school and eased the restrictions. No curfew was imposed during the pandemic and as of August 22, wearing face masks became compulsory only on public transport (16). Students received distance education for only 11 weeks. At the end of May, all grades returned to school with the warning to self-protect. The distance education has been led through Aula and EdTech Donor in Denmark (17). Aula is the common communication platform for staff, parents and students in primary schools and in day-care facilities. EdTech Donor is a website that provides a guide to different solutions that Denmark’s EdTech suppliers have made available to respond to COVID-19, with resources to support teaching, learning and training (17).

#### c) Survey and Data Collection

We conducted our research through a web-based survey created by the Survey Monkey software (SurveyMonkey, San Mateo, CA USA). The online survey contained two parts. The first part included a total of 54 questions that were related with socio-demographics, COVID-19, restrictions, distance learning and the psycho-social impact of the pandemic and lockdown. The second part included an evaluation of emotional status. We evaluated the negative and positive emotions of the subjects using Positive and Negative Affect Schedule (PANAS) (18) which was translated and validated into both Turkish and Danish (19, 20). The survey has been translated into Danish and has been sent to high school students at Copenhagen by Ms Ayumi Mayer.

#### d) Positive and Negative Affect Schedule

Emotional status was assessed by using the Positive and Negative Affect Schedule (PANAS), which consisted of 20 items, ten of which are used to measure positive affects (PA) and other ten to measure negative affects (NA). The schedule involves rating the effects on a Likert scale of 1 to 5 as to indicate the extent of how much they felt this effect: 1 through 5 corresponds to very slightly or not at all, a little, moderately, quite a bit, and extremely, respectively. The total score of each positive or negative effects category is obtained by adding all scores that would give a total score ranging between 10 and 50.

### Ethical statement

The study was conducted between July 3 and August 31. The study was approved by the Ministry of Health (08T14_39_03) and the Ministry of Education (59090411-20-E.10217175) in Turkey. Ethical committee of Cerrahpasa Medical School at Istanbul University-Cerrahpasa also approved the study (12/10/2020-134020). Electronic informed consent was presented on the first page of the survey citing that the survey is voluntary and participants could withdraw from the survey at any time.

### Statistical Analysis

Numeric results were shown as mean ± standard deviation, and categorical results were shown as number (percentage). Normality distribution of the numeric variables was tested by Shapiro Wilk test. The Mann Whitney U test was used for comparison of positive affects and negative affects by nationality (Turkish versus Danish), gender (male versus female), and whether students or family members/close acquaintances have COVID-19 (no versus yes). Those who identified themselves as ‘unidentified gender’ were excluded from the gender analysis because of the small numbers. Categorical data were compared by using the Chi-square test (Pearson, Yates, or Fisher Exact test). Relationships between positive affects and negative affects with gender, maternal and paternal education, type of school, grade, age, and income level were investigated using Spearman correlation coefficient. Reliability of the PANAS was assessed with Cronbach’s alpha coefficient. IBM SPSS Statistics for Windows, v.20.0 (IBM Corp., Armonk, NY, USA) was used in statistical analysis. P<0.05 was considered as statistically significant.

## RESULTS

### Demographic and socio-economic characteristics (Table 1)

A total of 565 (196 M/ 358 F/ 11 unidentified gender, mean age: 16.5 ± 1.0) Turkish students and a total of 92 (21 M/70 F/1 unidentified gender), mean age: 17.7 ± 1.0) Danish students were included in the study. In both countries, the majority of the respondents were females (63.4% Turkish/ 76.1% Danish). Among Turkish students, 12^th^ grade students were less likely to fulfill the questionnaire. Students educated in public (47.6%) and private high schools (52.4%) were nearly similar in number in Turkish group, whereas in the Danish sample almost all students were from public school (98.9%). Turkish parents were significantly less educated compared to Danish parents. We did not use purchasing power parity adjustment therefore the monthly income is not applicable for direct comparison, however, those who reported a higher monthly income were significantly less among the Turkish students. Moreover, those who reported family income loss during the pandemic were significantly higher among the Turkish students. The household number was < 5 in the majority among both Turkish and Danish students. Both Turkish and Danish students have filled up their free time mostly by using social networks, while, Turkish students have spent more time on it.

**Table 1.** Demographic and socio-economic characteristics

| n (%) | Turkish students, n= 565 | Danish students, n =92 | P |
| --- | --- | --- | --- |
| Gender |  |  | 0.059 |
| Male | 196 (34.7) | 21 (22.8) |  |
| Female | 358 (63.4) | 70 (76.1) |  |
| Undefined | 11 (1.9) | 1 (1.1) |  |
| Mean age ± SD | 16.52 ± 1.05 | 17.73 ± 1.09 | <0.001 |
| High school grade |  |  | NA |
| 9 | 172 (30.4) | 31 (33.7) |  |
| 10 | 152 (26.9) | 32 (34.8) |  |
| 11 | 171 (30.3) | 29 (31.5) |  |
| 12 | 70 (12.4) |  |  |
| Type of high school |  |  | <0.001 |
| Public | 269 (47.6) | 91 (98.9) |  |
| Private | 296 (52.4) | 1 (1.1) |  |
| Maternal educational status |  |  | <0.001 |
| University/higher education | 301 (53.3) | 71 (77.2) |  |
| High school/middle school | 172 (30.4) | 13 (14.1) |  |
| Primary school | 83 (14.7) | 7 (7.6) |  |
| Uneducated | 9 (1.6) | 1 (1.1) |  |
| Paternal educational status |  |  | 0.044 |
| University/higher education | 324 (57.4) | 67 (72.8) |  |
| High school/middle school | 177 (31.3) | 19 (20.7) |  |
| Primary school | 61 (10.8) | 6 (6.5) |  |
| Uneducated | 3 (0.5) | 0 (0.0) |  |
| Monthly household income |  |  | NA |
|  | Undefined:157 (27.8%)<br>>10.000 TL:145 (25.7%)<br>5000-10.000 TL:135 (23.9%)<br>2500-5000 TL:101 (17.9%)<br><2500 TL: 27 (4.8%) | Undefined/:25 (27.2%)<br>>80.000DKK:32 (34.8%)<br>40.000 - 80.000 DKK:21 (22.8%)<br>20.000 - 40.000 DKK:10 (10.9%)<br><20.000 DKK: 4 (4.4%) |  |
| Changes in family income during the pandemic |  |  | <0.001 |
| There has been loss | 162 (28.7) | 20 (21.7) |  |
| It did not change | 198 (35.0) | 56 (60.9) |  |
| I do not know | 205 (36.3) | 16 (17.4) |  |
| Household number |  |  |  |
| 1-4 | 421 (74.5) | 61 (66.3) | 0.150 |
| 5-7 | 140 (24.8) | 31 (33.7) |  |
| >7 | 4 (0.7) | 0 (0.0) |  |
| How did you spend your time (select all that apply)? |  |  |  |
| Social media | 484 (85.7) | 78 (84.8) | 0.824 |
| Education | 361 (63.9) | 72 (78.3) | 0.007 |
| Reading | 304(53.8) | 36 (39.1) | 0.009 |
| Sports | 283(51.0) | 40 (43.5) | 0.240 |
| Hobbies | 390(69.0) | 61 (66.3) | 0.602 |
| Family | 333(58.9) | 50 (54.4) | 0.408 |
| How long did you use social media (daily)? |  |  | 0.009 |
| <1 hour | 34 (6.0) | 12 (13.0) |  |
| 1-3 hour | 167 (29.6) | 30 (32.6) |  |
| >4 hour | 364 (65.4) | 50 (54.4) |  |

### COVID-19 knowledge, risk factors, restrictions, and precautions (Table 2)

The majority of the Turkish students declared that they were sufficiently well informed (73.1%) and were strongly following precautions (86.6%). Whereas these numbers were significantly lower among Danish students, with 59.8% of them reporting being sufficiently informed and only 15.2% adhering to the precaution at all times. Those who had a family member who had to leave the house during the pandemic at least a couple of times a week were significantly less among Turkish students (65.3%) compared to Danish students (80.5%). More Turkish students (10.8%) had an individual older than 65 years old at home than Danish students (7.6%). While none of the Danish students had contracted the virus themselves, 12% had reported that a family member or a close acquaintance had been diagnosed with the disease. On the other hand, a small group of Turkish students (0.4%) and their family members (2.7%) had contracted COVID-19.

**Table 2.** Questions about COVID-19 knowledge, risk factors, restrictions and precautions

| n (%) | Turkish students, n= 565 | Danish students, n =92 | P |
| --- | --- | --- | --- |
| How much information do you have about the COVID-19? |  |  | 0.001 |
| Sufficient | 413 (73.1) | 55 (59.8) |  |
| Moderate | 143 (25.3) | 33 (35.9) |  |
| Not enough | 3 (0.5) | 4 (4.4) |  |
| Not interested | 6 (1.1) | 0 (0.0) |  |
| How much did you follow the precautions (social distancing, washing hands, etc)? |  |  | <0.0001 |
| Always | 489 (86.6) | 14 (15.2) |  |
| Sometimes | 66 (11.7) | 75 (81.5) |  |
| Rarely | 6 (1.1) | 3 (3.3) |  |
| Never | 4 (0.7) | 0 (0.0) |  |
| Have you been diagnosed with COVID-19? |  |  | <0.001 |
| Yes | 2 (0.4) | 0 |  |
| No | 482 (85.3) | 92 (100.0) |  |
| I don't know | 81 (14.3) | 0 |  |
| Has any of your family members/or close acquaintances got diagnosed with COVID-19? |  |  | <0001 |
| Yes | 15 (2.7) | 11 (12.0) |  |
| No | 487 (86.3) | 81 (88.0) |  |
| I don't know | 62 (11.0) |  |  |
| How often have your family members left the house during the pandemic? |  |  | 0.012 |
| Almost everyday | 153 (27.1) | 34 (37.0) |  |
| A couple times a week | 216 (38.2) | 40 (43.5) |  |
| Rarely | 196 (34.7) | 18 (19.5) |  |
| Were there individuals older than 65 years at home? |  |  | 0.352 |
| Yes | 61 (10.8) | 7 (7.6) |  |
| No | 504 (89.2) | 85 (92.4) |  |

### Variables associated with distance learning (Table 3)

The majority of the students did not have difficulty in obtaining adequate technological equipment and study areas. While 73.6% of the Turkish students had their own technological devices and 13.8% have faced connection issues often, all of the Danish students had their own technological device, and only 8.7% have faced connection issues often. Turkish students had received less frequent online education compared to Danish students (daily schedule: 73.4% versus 91.3%). The demand for compensatory education to complete the second semester of the curriculum was significantly higher among Turkish students compared to Danish students (61.6% versus 18.5%). Despite these disparities across countries, online education created emotional stress and decreased the motivation similarly in the majority among both Turkish and Danish students. In both groups, approximately half of the students thought that their mental status was negatively affected by not going to school every day. Finally, the majority in both countries would have chosen face to face education if everything was to be normal (84.9% versus 95.6%).

**Table 3.**
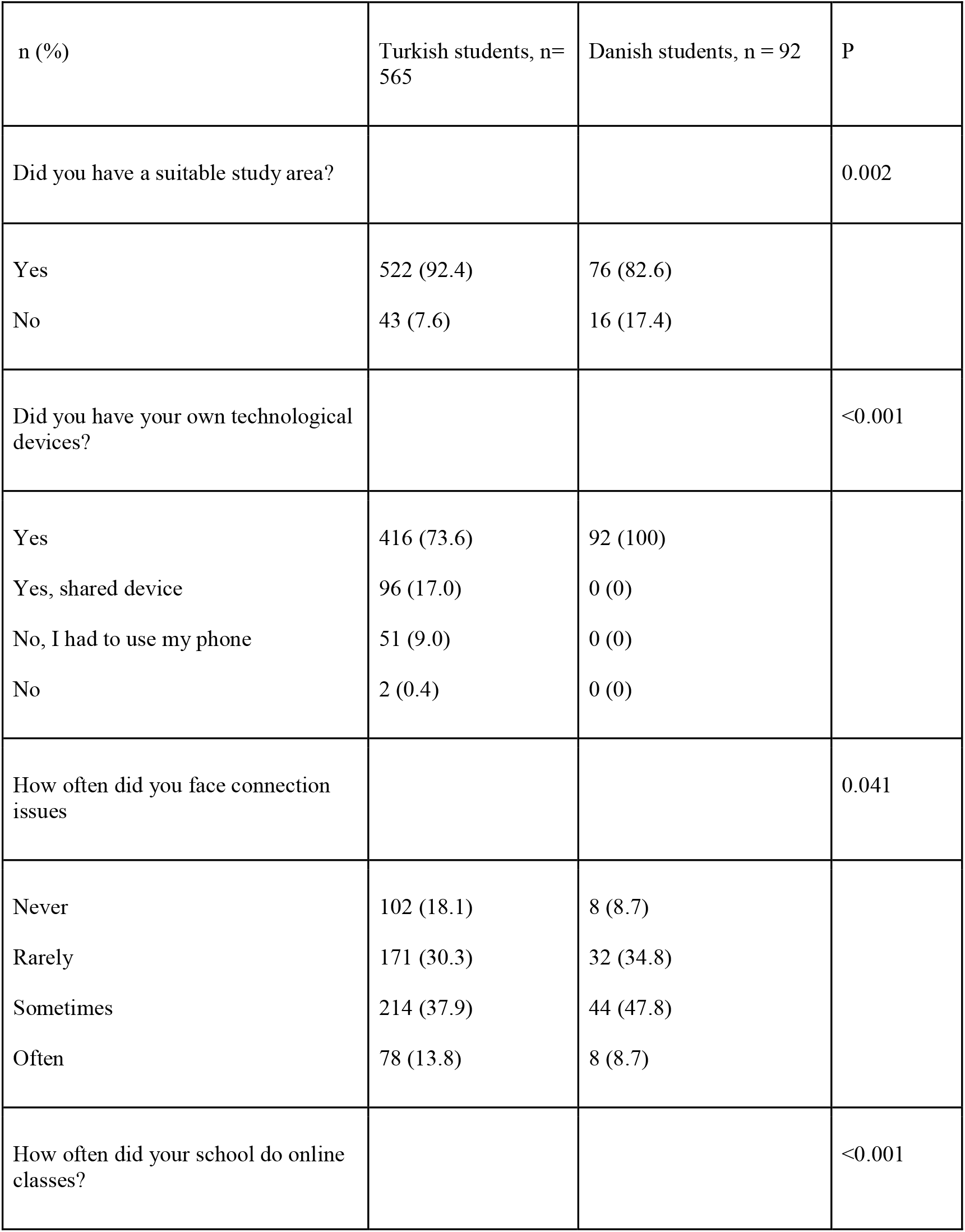
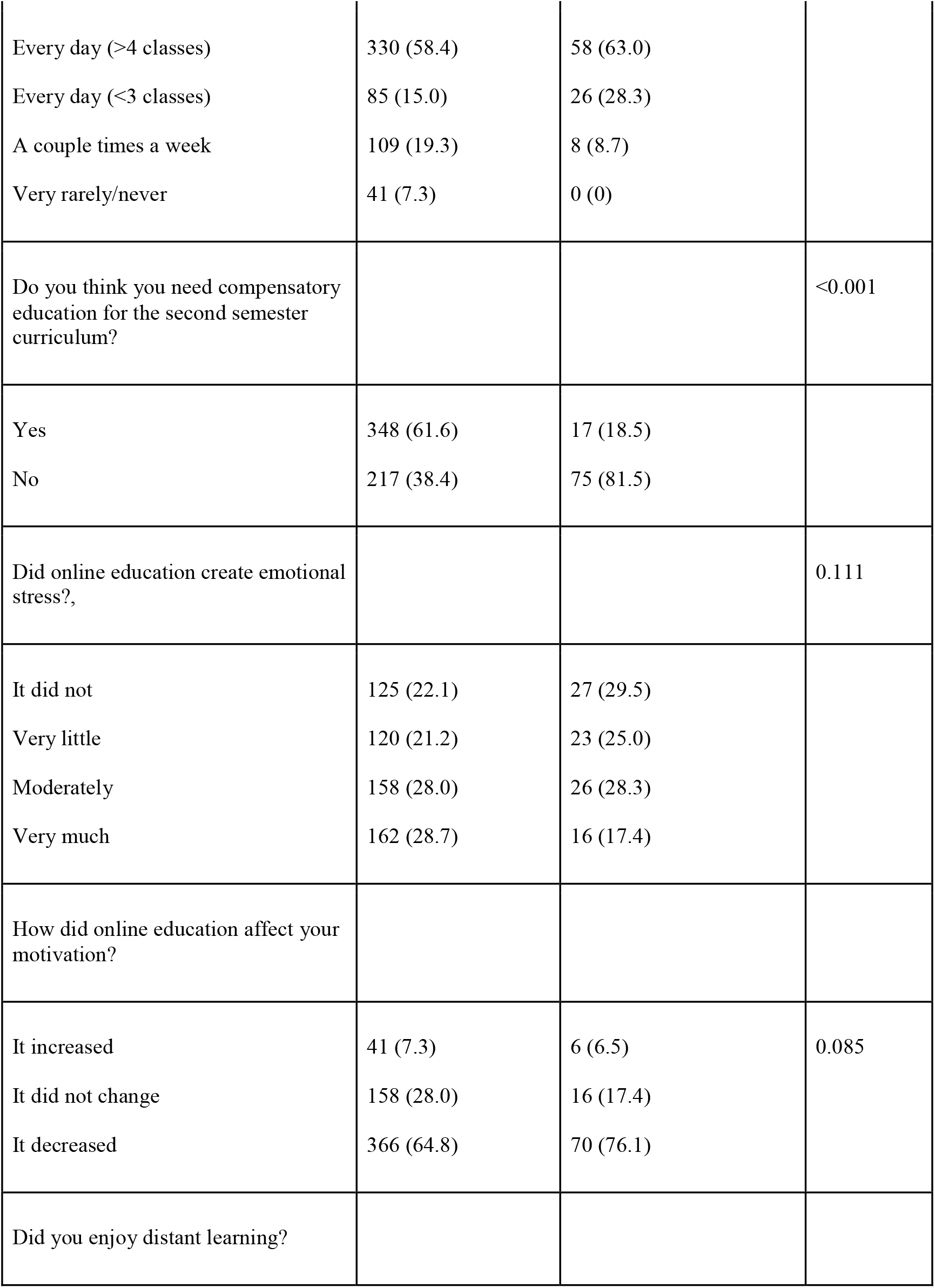
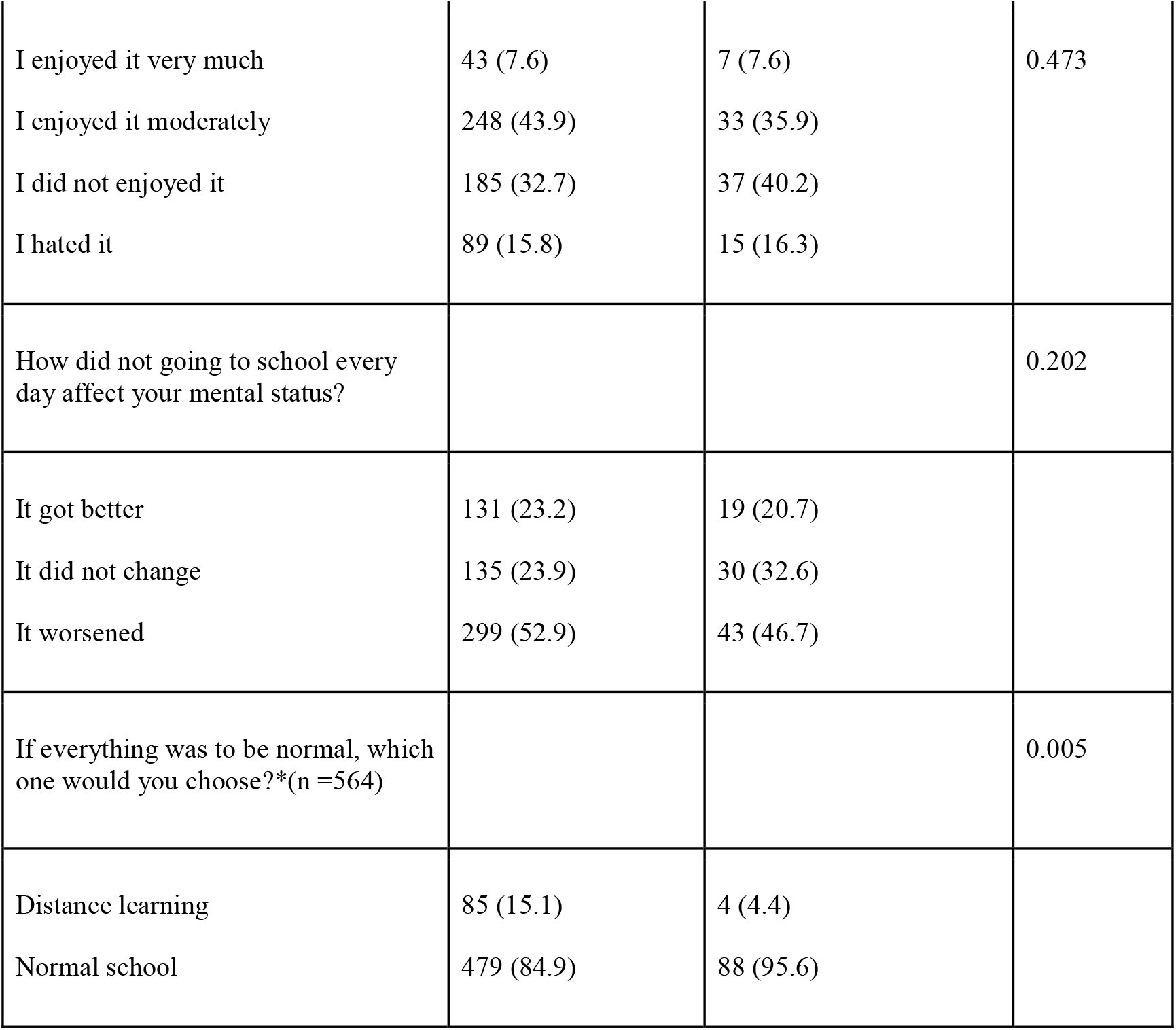
Technical problems, academical efficacy and psychological effects of distance learning

| n (%) | Turkish students, n= 565 | Danish students, n = 92 | P |
| --- | --- | --- | --- |
| Did you have a suitable study area? |  |  | 0.002 |
| Yes | 522 (92.4) | 76 (82.6) |  |
| No | 43 (7.6) | 16 (17.4) |  |
| Did you have your own technological devices? |  |  | <0.001 |
| Yes | 416 (73.6) | 92 (100) |  |
| Yes, shared device | 96 (17.0) | 0 (0) |  |
| No, I had to use my phone | 51 (9.0) | 0 (0) |  |
| No | 2 (0.4) | 0 (0) |  |
| How often did you face connection issues |  |  | 0.041 |
| Never | 102 (18.1) | 8 (8.7) |  |
| Rarely | 171 (30.3) | 32 (34.8) |  |
| Sometimes | 214 (37.9) | 44 (47.8) |  |
| Often | 78 (13.8) | 8 (8.7) |  |
| How often did your school do online classes? |  |  | <0.001 |
| Every day (>4 classes) | 330 (58.4) | 58 (63.0) |  |
| Every day (<3 classes) | 85 (15.0) | 26 (28.3) |  |
| A couple times a week | 109 (19.3) | 8 (8.7) |  |
| Very rarely/never | 41 (7.3) | 0 (0) |  |
| Do you think you need compensatory education for the second semester curriculum? |  |  | <0.001 |
| Yes | 348 (61.6) | 17 (18.5) |  |
| No | 217 (38.4) | 75 (81.5) |  |
| Did online education create emotional stress?, |  |  | 0.111 |
| It did not | 125 (22.1) | 27 (29.5) |  |
| Very little | 120 (21.2) | 23 (25.0) |  |
| Moderately | 158 (28.0) | 26 (28.3) |  |
| Very much | 162 (28.7) | 16 (17.4) |  |
| How did online education affect your motivation? |  |  |  |
| It increased | 41 (7.3) | 6 (6.5) | 0.085 |
| It did not change | 158 (28.0) | 16 (17.4) |  |
| It decreased | 366 (64.8) | 70 (76.1) |  |
| Did you enjoy distant learning? |  |  |  |
| I enjoyed it very much | 43 (7.6) | 7 (7.6) | 0.473 |
| I enjoyed it moderately | 248 (43.9) | 33 (35.9) |  |
| I did not enjoyed it | 185 (32.7) | 37 (40.2) |  |
| I hated it | 89 (15.8) | 15 (16.3) |  |
| How did not going to school every day affect your mental status? |  |  | 0.202 |
| It got better | 131 (23.2) | 19 (20.7) |  |
| It did not change | 135 (23.9) | 30 (32.6) |  |
| It worsened | 299 (52.9) | 43 (46.7) |  |
| If everything was to be normal, which one would you choose?*(n =564) |  |  | 0.005 |
| Distance learning | 85 (15.1) | 4 (4.4) |  |
| Normal school | 479 (84.9) | 88 (95.6) |  |

### Social and psychological effects of the COVID-19 pandemic or its restrictions (Table 4)

The majority among both Turkish and Danish students expressed loneliness, did not enjoy staying home, expressed a decrease in their physical activity and reported changes in their eating habits. Moreover, sleep disturbances were observed in about half of the students in both groups. Turkish students seemed to be more worried about their plans ‘to study abroad’. Similarly, the feelings of “boredom of life”, “anxiety towards the future” and the idea of “becoming more mature with the pandemic” were significantly more common among Turkish students. Students in both countries thought that in the long term the economy and social life would be affected by the pandemic, while this was more pronounced among the Danish students. Finally, both Turkish and Danish students reported domestic physical (9.9% versus 9.8) and emotional abuse (36.3 versus 37.0%).

**Table 4.** The social and psychological effects of the COVID-19 pandemic or its restrictions

| n (%) | Turkish students, n=565 | Danish students, n=92 | P |
| --- | --- | --- | --- |
| Did your plans get affected if you are planning on studying abroad? |  |  | 0.040 |
| Yes | 124 (21.9) | 11 (12.0) |  |
| No | 149 (26.4) | 33 (35.9) |  |
| I am not thinking of studying abroad | 292 (51.7) | 48 (52.2) |  |
| In the future do you think the pandemic will still affect the economy? |  |  | 0.016 |
| Yes | 164 (29.0) | 34 (37.0) |  |
| No | 237 (42.0) | 24 (26.0) |  |
| Undecided | 164 (29.0) | 34 (37.0) |  |
| What do you think about the long-term social effects of the pandemic? |  |  | 0.181 |
| It scares me/ makes me anxious | 272 (48.1) | 50 (54.4) |  |
| I think that the world will be a better place | 130 (23.0) | 24 (26.1) |  |
| I don't think that it will affect a lot | 163 (28.9) | 18 (19.5) |  |
| How did your anxiety about your future get affected? |  |  | <0.001 |
| It increased | 347 (61.4) | 21 (22.8) |  |
| It did not change | 204 (36.1) | 67 (72.8) |  |
| It decreased | 14 (2.5) | 4 (4.4) |  |
| How did your feeling of loneliness get affected? |  |  | 0.706 |
| It felt less lonely | 37 (6.6) | 5 (5.4) |  |
| It did not change | 216 (38.2) | 32 (34.8) |  |
| I felt more lonely | 312 (55.2) | 55 (59.8) |  |
| How did your boredom of life get affected? |  |  | 0.019 |
| It got better | 42 (7.4) | 6 (6.5) |  |
| It did not change | 121 (21.4) | 32 (34.8) |  |
| It worsened | 402 (71.2) | 54 (58.7) |  |
| Did the pandemic make you more mature /helped you grow? |  |  | <0.001 |
| Yes | 260 (46.0) | 22 (23.9) |  |
| No | 211 (37.4) | 44 (47.8) |  |
| Undecided | 94 (16.6) | 26 (28.3) |  |
| Did you enjoy spending time at home during the pandemic? |  |  | 0.063 |
| Yes | 185 (32.7) | 32 (34.8) |  |
| No | 198 (35.1) | 41 (44.6) |  |
| Undecided | 182 (32.2) | 19 (20.6) |  |
| How did your physical activity get affected? |  |  | 0.296 |
| It increased | 112 (19.8) | 24 (26.1) |  |
| It didn't change | 120 (21.2) | 21 (22.8) |  |
| It decreased | 333 (59.0) | 47 (51.1) |  |
| How did your eating habits get affected? |  |  | 0.193 |
| I started eating more/irregularly | 238 (42.1) | 40 (43.5) |  |
| It didn't change | 170 (30.1) | 32 (34.8) |  |
| I started eating less | 157 (27.8) | 20 (21.7) |  |
| Did you have problems with falling asleep? |  |  | 0.313 |
| Very troubled | 173 (30.6) | 21 (22.8) |  |
| Little troubled | 153 (27.1) | 30 (32.6) |  |
| It didn't change | 195 (34.5) | 36 (39.1) |  |
| I started to sleep easier | 44 (7.8) | 5 (5.4) |  |
| How often did your sleep get divided? |  |  | 0.269 |
| Very often | 99 (17.5) | 10 (10.9) |  |
| Sometimes | 219 (38.8) | 40 (43.5) |  |
| Never | 247 (43.7) | 42 (45.6) |  |
| Did the domestic physical abuse at home increase? |  |  | 0.005 |
| Yes | 56 (9.9) | 9 (9.8) |  |
| No | 476 (84.3) | 69 (75.0) |  |
| Undecided | 33 (5.8) | 14 (15.2) |  |
| Did the domestic emotional abuse at home increase? |  |  | 0.582 |
| Yes | 205 (36.3) | 34 (37.0) |  |
| No | 300 (53.1) | 45 (48.9) |  |
| Undecided | 60 (10.6) | 13 (14.1) |  |

### PANAS scores (Table 5)

The majority of the students who participated in the survey had completely fulfilled the PANAS scale (Turkish sample: 89%, 503: 168 M/ 326 F/ 9 undefined gender; Danish sample: 79%, 73: 18 M/ 54 F/ 1 undefined gender). Cronbach’s alpha coefficients of the PANAS were calculated as 0.81 for negative subscale and 0.82 for positive subscale indicating good reliability.

**Table 5.** Comparison of positive and negative affects

|  |  | Turkish students<br>(503: 168 M/ 326 F/ 9 undefined gender*) | Danish students<br>(73: 18 M/ 54 F/ 1 undefined gender*) | P |
| --- | --- | --- | --- | --- |
| Total | Positive Affects | 26.95 ± 7.59 | 25.82 ± 5.63 | 0.217 |
|  | Negative Affects | 24.84 ± 7.52 | 24.45 ± 7.32 | 0.742 |
| Male | Positive Affects | 29.11 ± 7.61† | 25.00 ± 5.78§ | 0.026 |
|  | Negative Affects | 22.21 ± 6.96‡ | 22.67 ± 7.28¶ | 0.827 |
| Female | Positive Affects | 25.92 ± 7.33† | 26.13 ± 5.65§ | 0.767 |
|  | Negative Affects | 26.20 ± 7.38‡ | 25.00 ± 7.37¶ | 0.293 |

\*: Those who identified themselves as ‘unidentified gender’ were included in total calculations of positive and negative affects, however, were excluded from the gender analysis
†: Turkish students: male vs female positive affects: p <0.001
‡: Turkish students: male vs female negative affects: p <0.001
§: Danish students: male vs female positive affects: p =0.575
¶: Danish students: male vs female negative affects: =0.206

The total scores of PANAS were similar between Turkish and Danish students. When countries were separately analyzed, the total score of positive affects was significantly higher among males (p<0.01), whereas that of the negative score was significantly higher among females (p<0.01) in the Turkish group (Table 5). On the other hand, there were no such gender differences in the Danish group. When males and females were separately analyzed, positive affects score was calculated significantly higher among Turkish male students compared to Danish counterparts, besides that, no significant difference across two countries was observed.

We made a brief literature review of studies investigating PANAS in the adolescent population and presented their main results in Table 6 (21-25). All studies were done among apparently healthy adolescents before the COVID-19 pandemic. Compared to the previous studies, in our study, total PA values were lower whereas NA values were higher.

**Table 6.** Review of the literature related with PANAS in adolescents

| Reference No | First author | Year of publication | Country | Participants, n (Male, %) | Age, years | PA | NA |
| --- | --- | --- | --- | --- | --- | --- | --- |
| (21) | Roberts K.R. | 1998 | USA | 126 (36) | 15.7±2.2 | 33.2±8.6 | 14.6±7.7 |
| (22) | Allan N.P. | 2015 | USA | 608 (52.3) | 15.45±1.09 | 35.26±10.46 | 20.02±8.86 |
| (23) | Ortuño-Sierra J. | 2019 | Spain | 1032 (51.5) | 11.91±1.37 | 38.46±6.13 | 22.91±6.82 |
| (24) | Staes F | 2003 | Belgium | 620 (46.9) | 17.1 ±0.68 (back pain)<br>17.1 ±0.64 (no back pain) | 35 (32-37) (back pain)<br>35 (31-38)(no back pain) | 22 (17-27) (back pain)<br>20 (16-23) (no back pain) |
| (25) | Heubeck B.G. | 2020 | Australia | 1431 (100) | 14.62±1.73 | 35.06±6.21 | 20.84±6.81 |
| Current study | Seyahi LS | Not defined | Turkey<br>Denmark | 565(34.7)<br>92 (22.8) | 16.52 ± 1.05<br>17.73 ± 1.09 | 26.95 ± 7.59<br>25.82 ± 5.63 | 24.84 ± 7.52<br>24.45 ± 7.32 |

Data are expressed as mean±SD
Positive and Negative Affect Schedule (PANAS).
PA: Positive affects; NA: Negative affects.

### Variables associated with PANAS

Being female (only Turkish group), the knowledge about COVID-19 (only Turkish group), having been diagnosed as COVID-19 or having a friend or a family relative infected with COVID-19 (only Danish group), increased daily social media use (only Turkish group) and lower income (only Turkish group) were found to be associated with PANAS scores (data not shown).

### Comparison between public and private school students in Turkey

The frequency of female students was similar in public and private schools (public: 61.0%, private: 65.5%). Parental education level and monthly family income were significantly lower among public school students. Public school students were less likely to have adequate study room, private technological devices and undisrupted internet connection compared to private school students (data not shown). Among public school students, the frequency of online classes was significantly less (daily schedule: 53.1% versus 91.9%, p<0.001), the need for supplementary education was significantly higher (66.5% versus 57.1, p =0.021) and the frequency of those who do not think that they completed the second half of the curriculum was significantly higher (75.5% versus 57.1%, p<0001) compared to private school students. Both public and private school students found EBA broadcasts of the Turkish Ministry of Education ineffective (89.1% versus 91.6%). Similarly, the knowledge about COVID-19, the compliance with the restrictions, dissatisfaction from distance education and the psychological effects of the pandemic including sleeping and eating problems were similar between those who were educated in public and private schools (data not shown). Furthermore, the PANAS values did not differ between public and private school students (PA: 26.8 ± 7.5 versus 27.1 ± 7.6, p=0.570; NA: 24.7± 7.2 versus 25.0± 7.8, p=0.598).

## DISCUSSION

In this cross-sectional web-based survey done among high school students from two different countries during the summer of 2020 while the COVID-19 outbreak was still going on, we observed that social and psychological status of the adolescents were heavily affected by the pandemic and that the students were discontent with the distance education and would prefer face to face education when everything returns to normal. These observations were true regardless of the socio-economic differences of the students across Turkey and Denmark as well as within Turkey across public and private schools.

The high school students reported increased feelings of “loneliness” and “boredom”, as well as heightened levels of ‘anxiety about the future effects of the ongoing COVID-19 pandemic’ similar to what have been observed in previous studies (2, 4-10, 26-30). They also reported lower levels of positive affects, and higher levels of negative counterparts, compared to the previous studies investigating the adolescents prior to the pandemic (21-25). These findings indicate a worsening emotional status in the high school students, as a consequence of the ongoing COVID-19 pandemic.

A significant proportion of the students among both countries reported that after the onset of the pandemic, their physical activity level decreased (59.0% versus 51.1%), and that they started eating more or irregularly (42.1% versus 43.5%). These indicate that the pandemic not only has affected the emotional status of students, but also altered the contents of the daily activities in line with what have been observed in previous studies (31-33). Moreover, a noteworthy percent of the students (22% versus 30%) reported severe trouble falling asleep and fragmented sleep (10% versus 17%). Very recently, the prevalence of insomnia symptoms during the epidemic was found to be 23.2% among 11,835 adolescents and young adults (34). Insomnia symptoms were associated with depression and anxiety and seemed to be relieved with social support (34). Furthermore, isolation, obligatory stay at home and withdrawal from social life may lead to increased sedentary behaviors and food consumption which could affect sleep (32). It has been also suggested that children and adolescents could be also influenced by the changes in family financial situations, health concerns, and uncertainty about the future (32). Reduced exposure to sunlight as a result of pandemic related restrictions may disturb the sleep routine (32). Finally, challenges in distance learning, decreased educational motivation, absence of face to face contact and excess use of social media could be further factors that may lead to sleep problems (32).

It has been suggested that based on previous pandemics experience, there is an increasing concern about increasing domestic violence (35, 36). Also international organizations, social media and the communication broadcasts around the world have expressed their growing concern on this matter; however formal data about this issue is scarce. In our study we found that over one-third of the students among both countries reported an increase in domestic emotional abuse, and one-tenth complained of increased domestic physical abuse. This raises concerns about the home environment, becoming an unstable and, probably, a traumatic one during the pandemic, which can severely impact some students’ emotional well-being by acting as an emotional stressor.

In our study, more Turkish students compared to the Danish counterparts had concerns about their future affected badly by the pandemic (61.4% versus 22.8%), felt that their future plans on studying abroad could be influenced (21.9% versus 12.0%), and had worsening feelings of “boredom of life” (71.2% versus 58.7%). Also, significantly more Turkish students than the Danish ones reported that they complied with the precautions (86.6% versus 15.2%), and that they had sufficient information about the COVID-19 (73.1% versus 59.8%). These differences between the Turkish and the Danish students might be due to the socio-cultural and economic differences between the two populations, as well as the significant differences between the pandemic related measures taken by the two countries as described earlier (see Methods). Social measures like lockdowns are life-changing events, and particularly when prolonged, might have behavioral and emotional consequences on the affected populations.

We observed that there were significant socio-economic disparities defined as parental education and monthly income between Turkish and Danish students and also within Turkey between public and private school students. In line with that, Turkish students were less likely to have private technological equipment and faced connection problems significantly more frequently than Danish students. Furthermore, Turkish online education system was significantly less adequate and satisfactory compared to the Danish system implied by the less frequent online classes and high demand for future compensatory education. In fact, using income, parental education level, technological resources and qualified online education, we were able to stratify all study participants into 3 groups: while Turkish students who were attending public schools constituted the most economically disadvantaged group, Danish students were the least. It has to be noted that except one, all Danish students were educated in public schools, indicating the high standards of the social system in Denmark. These observations indicate a large opportunity gap between students which was probably present before the pandemic yet becoming wider with school closures. According to the Turkish Ministry of Education 2017-2018 academic year report, the rate of private high school students among all high school students is 10.4% (37). Therefore one can assume that currently, great majority of the high school students in Turkey do not receive an effective education. Of note, face to face education has not been started yet, as of October 18, 2020, while this manuscript was being prepared for submission. Moreover, regardless of the unequal opportunities, we observed that the great majority of the students in both countries disapproved distance learning (3, 38-40). Similar to our results, several studies reported that the distance learning was not as effective as normal education, might decrease their motivation and can cause anger and frustration (3, 38-40).

Study participants in our study were mostly girls, which was true for both countries. This was similar to what has been observed in previous surveys investigating psychological status during the Covid-19 pandemic (4, 5, 41, 42). In line with our observations, being female, having a lower level of education and income, excess social media use and having been diagnosed with COVID-19, were found to be associated with psychological status (2, 4, 9, 27, 29-31). In our study, PANAS scores of Turkish students indicated that females compared to males were more severely affected. On the other hand, such gender difference was not found in the Danish group. Among female students, both PA and NA scores were similar between Turkish and Danish students. On the other hand, among males, while, NA scores were similar, PA scores were significantly lower among Danish students. While this could be due to the small sample size of the male students in the Danish group, it could be also due to the fact that Danish male students might have been as sensitive as their female counterparts, indicating a cross cultural difference.

Our study has several limitations. Information bias and lack of longitudinal data are inherent to cross-sectional questionnaire study design. Oversimplification of reality is also one of the limitations of the questionnaire study design because of the multiple-choice questions with preconceived categories. It should be noted that socio-cultural status might also affect an individual’s responses. Also, the size of the Danish group was small (1/5^th^ of the Turkish group) which could affect statistical calculations. Especially, the male group within the Danish sample was small and this might cause type 2 error. The effect of social isolation and distance learning might be correlated with the duration. Finally, despite the short duration of data collection, our study groups might not be homogeneous in terms of this effect size. For some time Turkish students have been having distance education while Danish students have already started their semester in class. This fact can cause discrepancies between the Danish survey and Turkish survey outputs.

In conclusions, we found that both COVID-19 pandemic and distance education had negative effects on mood statues among both Turkish and Danish students. Students reported lower levels of positive affects, and higher levels of negative counterparts, compared to the previous studies done prior to the pandemic. Female students were significantly more severely affected; however this was true for only the Turkish group. Moreover, students expressed loneliness, boredom and anxiety towards the future. Decreased physical activity, sleep problems, eating disorders and domestic abuse were other serious complaints. Despite the fact that there were various socio-economic differences between two countries, the psychological impact was almost comparable between Turkish and Danish students. Turkish online education system especially that taught by the public schools was not found to be effective. Finally, the great majority in both countries expressed negative opinion about distance education. Considering the pandemic and its related restrictions could continue for an indefinite period of time, efficient social welfare measures should be taken by the states and international organizations in order to mitigate its adverse effects.

## Data Availability

All data referred in the manuscript are available upon request.

